# Silence in physician clinical practice: a scoping review protocol

**DOI:** 10.1101/2024.07.10.24310250

**Authors:** Martina Ann Kelly, Stefanie Rivera, Caitlin McClurg, Catherine Sweeney, Stephen Mosca, Ellen McLeod, Deirdre Bennett, Megan Brown

**Affiliations:** Department of Family Medicine, Cumming School of Medicine, University of Calgary, Calgary, Alberta, Canada; Libraries and Cultural Resources, University of Calgary, Calgary, Alberta, Canada; Medical Education Unit, Brookfield Health Sciences Complex, University College Cork, Cork, Ireland; Division of Palliative Care, University of British Columbia, Canada; Division of Palliative Care, University of Calgary, Calgary, Alberta, Canada; School of Medicine, Newcastle University, Newcastle, United Kingdom

**Author notes:** **Corresponding author:** Catherine Sweeney, **Address:** School of Medicine, Brookfield Health Sciences Complex, University College Cork, Cork, Ireland, **Email:**.

**Keywords:** silence, communication, non-verbal communication, scoping review

## Abstract

**Objective:** The objective of this review is to map, describe and conceptualize how silence is discussed within literature on interactions between physicians and patients, in clinical settings.

**Methods:** We will use the methodological framework of Arksey & O’Malley, adapted by Levac et al and Joanna Briggs Institute. Empirical studies including quantitative, qualitative, mixed methods, observational studies and reviews will be included. Commentaries, editorials, and grey literature will also be examined. The databases MEDLINE, Cumulative Index to Nursing and Allied Health Literature, PsycINFO, Scopus and Web of Science will be searched. A two-part study selection strategy will be applied. First, reviewers will follow inclusion and exclusion criteria based on ‘Population-Concept-Context’ framework to independently screen titles and abstracts. Next, full texts will be screened. Data will be extracted, collated, and charted to summarize methods, outcomes and key findings from the articles included.

**Expected results and implications:** This scoping review will provide an extensive description of how physicians engage with silence in clinical settings. Findings will identify how silence is perceived in physician patient interactions, the roles it plays, what factors influence use of silence and guide development of educational initiatives on use of silence in clinical settings.

## Introduction

Effective use, and interpretation, of silence is a sophisticated and essential communication skill.^1^ Proficient use of silence is associated with enhanced empathy, understanding, thoughtfulness, and self-awareness,^1, 2^ essential for good medical practice.^3, 4^ Despite this, research into silence in healthcare communication is lacking and the topic is poorly covered in communication skills curricula.^5, 6^ Having an in-depth understanding of physicians’ use, and experiences of, silence, could apprise communication skills training to enhance good doctor-patient communication.

Silence is not merely the absence of verbal communication, but rather silence and speech organize each other, forming a continuum.^7, 8^ Silence in clinical contexts is defined as an absence of verbal audio signal, lasting longer than required to take turns to speak (2 seconds).^9^ The meaning of silence is impacted by the context, ambient sounds, utterances before and after the silence. Silence is also configured through non-verbal cues such as eye contact, gestures, movement, posture, and paralinguistic communication.^10–12^ Culture also plays a role in silence. For example, in Asian contexts silence can be a sign of respect and is also acknowledged as being full of meaning. In contrast, in Western cultures, especially North America, silence can be perceived more negatively, as a sign of unfriendliness or not being worthy – we must ‘add’ something to the conversation.^13^

Silence can play many roles in a consultation. In day-to-day language, silence, as expressed through pauses, is used to organize speech, such as turn-taking. For example, a physician may pause to invite a response, giving a participant a moment to gather their thoughts and think a bit longer about the subject matter. Silence can also afford emotional acknowledgement, for example following a moment of gravity. Silence can be non-productive, as in awkward silences, when the information being communicated is ambiguous or poorly understood. An awkward silence can also arise in the context of uncertainty, or distraction/ inattention. Non-productive silence can also be hostile, as in the withholding of information or judgement – a ‘conspiracy of silence’, or to prevent the sharing of anxiety. To date, several authors have attempted to characterize silence, using different terms such as compassionate silence,^14, 15^ connectional silence,^9, 15^ profound silence^16^ and awkward silence.^14^

Despite its significance, silence as a focus of research in healthcare is relatively limited, primarily originating from psychotherapy or palliative care.^14, 17^ A 2008 meta-ethnography of silence identified 18 studies, of which only 4 were empirical studies, the remainder consisting of opinion pieces, or commentaries.^17^ This review identified that studies drew on literature from psychology, communication, and spiritual traditions. More recently, researchers in oncology and palliative care have audio recorded consultations documenting the epidemiology of silence, coding for frequency of silence, duration, and several authors propose varying typologies of silence, often related to the duration and purpose of the pause.^9, 14, 18, 19^ However, the relationship between silence, nonverbal cues, and verbal communication, and how they influence each other, remains unclear. This scoping review seeks to address this research gap by examining how silence is described and conceptualized in the clinical literature. This protocol delineates the procedures for conducting the review, guided by good practice and protocols for scoping review development.^20^

The primary objective of this review is to identify, analyze and synthesize how silence is engaged in interactions between physicians (including physicians in training) and patients, in the clinical setting. This information will, we anticipate, be useful to enhance doctor-patient interactions through communication skills training.

Review questions:

1. How is silence conceptualized in the clinical literature involving doctors and patients?
2. What roles / functions does silence play in physician-patient communication?

## Methods

We chose to conduct a scoping review given the breadth of ways in which silence can be engaged. Scoping reviews are well suited to answer broad and exploratory research questions. They are used to explore new research areas, to clarify key concepts and identify research gaps by mapping the literary landscape, elucidating methodologies, core concepts, evidence types, and characteristics.^21^They frequently unveil a wider spectrum of evidence, serving as a foundation for systematic reviews and pinpointing knowledge voids.^20, 21^ This protocol has been reported using the Preferred Reporting Items for Systematic Reviews and Meta-analyses extension for systematic review protocols (PRISMA-P)^22^ (S1 Appendix).

## Methodological Framework

Our scoping review will follow the Arksey and O’Malley framework for scoping reviews,^23^ adapted by Levac et al.^24^ and the Joanna Briggs Institute.^25^ Components will include: identifying a research question; identifying relevant studies; study selection; charting data; collating, summarizing, and reporting results; and consultation. The findings of the review will be presented following the guidelines of the Preferred Reporting Items for Systematic Reviews and Meta-analyses extension for scoping reviews (PRISMA-ScR).^26^

## Stage 1: Identifying the research question

We used the Population Concept Context framework (PCC), recommended by the Joanna Briggs Institute for scoping reviews to develop our review question. ^25^ The population are physicians and patients, the concept is the experience of silence in professional caregiving interactions with physicians and patients, and the context is clinical settings. Our review question is ‘how is silence conceptualized in clinical literature involving doctors and patients?’ This question may be refined, or new ones added, as the authors gain increasing familiarity with the literature.

## Stage 2 Identifying relevant studies

*Types of Sources.* Empirical research on silence encompassing various study designs will be considered, including qualitative studies, observational studies, surveys and questionnaires, longitudinal studies, meta-analyses or evidence synthesis, and conversational analysis. We will also include commentaries, personal reflections and grey literature sources (conference proceedings, abstracts, thesis etc). Only English-language sources will be included due to feasibility and translation issues, with no restrictions on publication dates.

A preliminary search was conducted on Google Scholar to gain an overview of existing literature and identify seed studies. Text word from titles, and abstracts of seed papers, along with the MeSH terms from MEDLINE were tailored to develop an initial search strategy for MEDLINE (table 1). Comprehensive searches will be carried out in the following databases: Scopus, Web of Science, CINAHL Plus with Full Text, APA PsychINFO, and MEDLINE. Reference lists of included studies will undergo screening to identify any additional relevant studies. The search strategy will be adapted for each database and further refined in consultation with a research librarian (CmC).

**Table 1.** Inclusion and exclusion criteria according to the PCC framework.

|  | Inclusion criteria | Exclusion criteria |
| --- | --- | --- |
| Population | Physicians, ranging from general | Other healthcare professionals, such |
|  | practitioners to specialists, resident physicians, and medical students. | as Professional Nurses, Registered Nurses, Enrolled Nurses, Nurse Aides, Nursing Students, Dentists, Pharmacists, Nutritionists, EMTs, and Medical Laboratory Technologists. Additionally, Professional Therapists, Clinical Psychologists, Counseling Psychologists, as well as Professional Pastors and Spiritual Counselors |
| Concept | The role of silences in professional caregiving interactions with patients and clients.<br>This involves analyzing how silence is used in clinical settings and communication skills curricula.<br>These roles encompass silence in communication, verbal and non-verbal, as well as the concept of the "conspiracy of silence," where silence is wielded as a tool for power or control | Silence in religious contexts<br>Silence-external to human interaction<br>e.g. nature<br>Silence in relation to hearing impairment |
| Context | Clinical settings, such as hospitals, | Academic environments such |
|  | clinics, physician's offices, urgent care centers, nursing homes, long-term care facilities, rehabilitation centers, mental health clinics, palliative care units, and home care environments<br>Telemedicine | classroom, simulations, lectures etc. |

**Table 2.** Search strategy terms for MEDLINE (PubMed)

|  | MeSH terms | Related terms |
| --- | --- | --- |
| Population | exp Education,<br>Medical/ | provider*.tw,kf.<br>clinician*.tw,kf. |
|  | exp "Internship and Residency"/<br>exp Faculty, Medical/<br>exp Physician-Patient Relations/ | trainee*.tw,kf.<br>residen*.tw,kf.<br>interns*.tw,kf.<br>preceptor*.tw,kf.<br>physician*.tw,kf.<br>doctor*.tw,kf. |
| Concept | exp Communication<br>exp Mindfulness<br>exp Voice<br>exp Social Isolation/<br>or exp Loneliness/<br>exp Interpersonal Relations | silence*.tw,kf.<br>discuss*.tw,kf. ,support*.tw,kf., relation*.tw,kf.<br>connect*.tw,kf., communicat*.tw,kf., convers*.tw,kf.,<br>interact*.tw,kf., contemplat*.tw,kf., acknowledg*.tw,kf.<br>peace*.tw,kf., hope*.tw,kf.<br>paus*.tw,kf., mindful*.tw,kf.<br>awkward*.tw,kf., grie*.tw,kf.<br>stigma*.tw,kf., violen*.tw,kf., abus*.tw,kf<br>solitude.tw,kf., still*.tw,kf.,tranquil*.tw,kf |
| Context | exp Telemedicine/<br>exp Videoconferencing/<br>exp Remote Consultation/ | (telehealth* or teleconsult* or teleconf* or virtual care or zoom* or skype*).mp |

## Stage 3: Study Selection

After completing the search, all located records will be uploaded to Covidence systemic review software (Veritas Health Innovation) and duplicates removed. A pilot test will be performed on a random subset of 50 titles/abstracts to refine the inclusion/exclusion criteria, if necessary, and to ensure consistent application of selection criteria among reviewers.

Screening will take place in 2 phases. All citations will be screened independently by 2 reviewers, based on title and abstract. Any discrepancies that arise during each stage of the selection process will be resolved by consulting a third reviewer. Next full texts will be imported into Covidence and reviewed by 2 independent reviewers. Reasons for exclusion will be documented. Findings of the search and the study inclusion process will be comprehensively reported in the final scoping review, following the reporting guidelines outlined in the Preferred Reporting Items for Systematic Reviews and Meta-analyses Extension for Scoping Reviews (PRISMA-ScR).^26^

## Stage 4: Charting the data

Information drawn from each publication will encompass details such as authorship and year of publication, location, and study methodology. Additionally, particulars about the study population, concept, context, and pertinent findings relevant to the review question. A preliminary version of the data extraction tool is provided in Table 3.

**Table 3.**
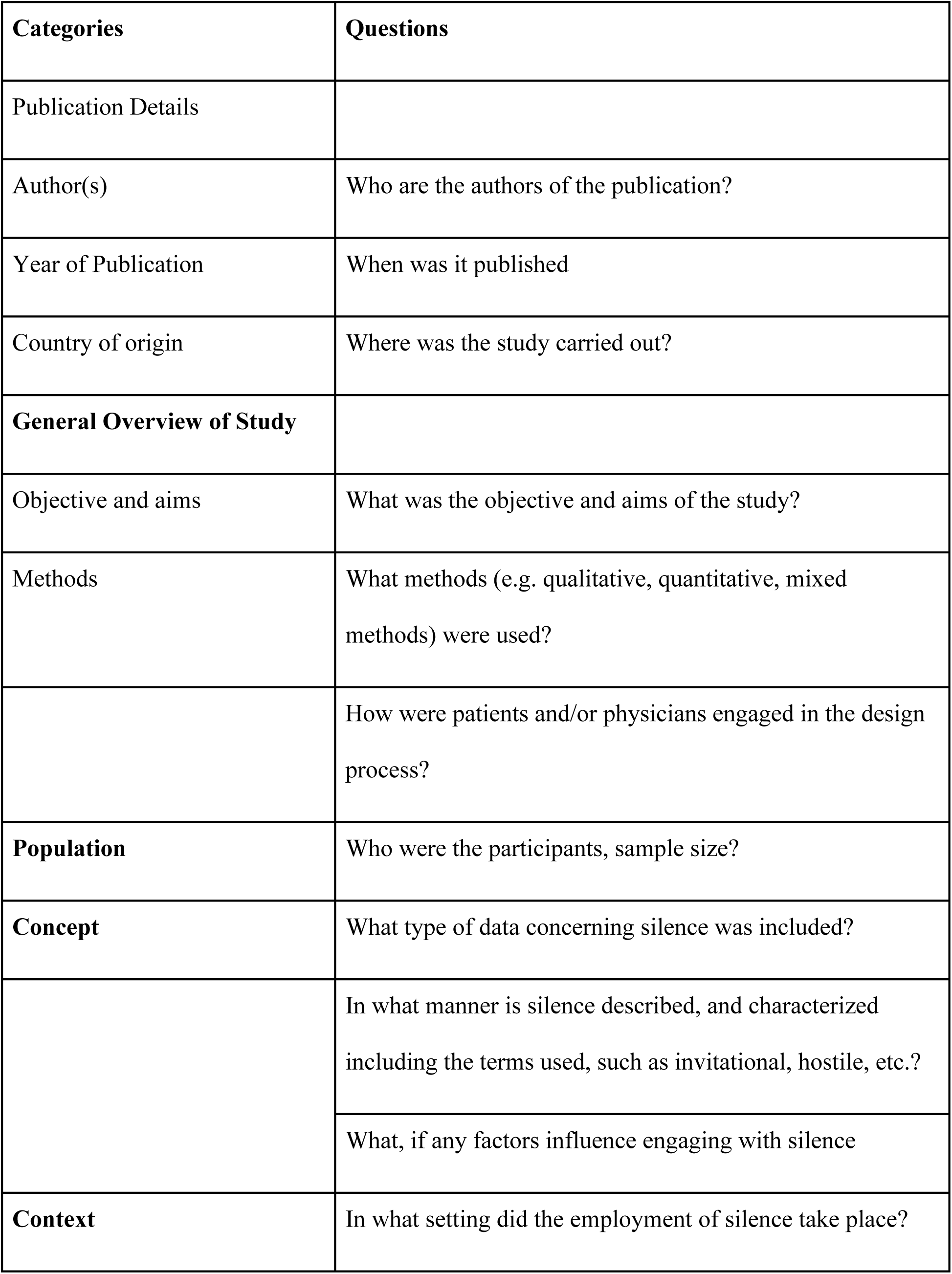
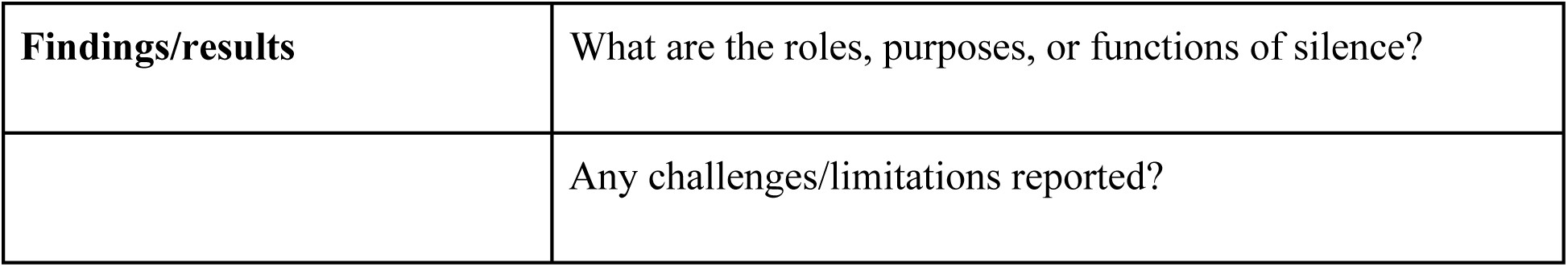
Preliminary Data Extraction Tool.

| **Categories** | **Questions** |
| --- | --- |
| Publication Details |  |
| Author(s) | Who are the authors of the publication? |
| Year of Publication | When was it published |
| Country of origin | Where was the study carried out? |
| **General Overview of Study** |  |
| Objective and aims | What was the objective and aims of the study? |
| Methods | What methods (e.g. qualitative, quantitative, mixed methods) were used? |
|  | How were patients and/or physicians engaged in the design process? |
| **Population** | Who were the participants, sample size? |
| **Concept** | What type of data concerning silence was included? |
|  | In what manner is silence described, and characterized including the terms used, such as invitational, hostile, etc.? |
|  | What, if any factors influence engaging with silence |
| **Context** | In what setting did the employment of silence take place? |
| **Findings/results** | What are the roles, purposes, or functions of silence? |
|  | Any challenges/limitations reported? |

Before the extraction process, the team will collaborate to pilot five of the included studies to confirm accuracy, ensure mutual comprehension, and assess the suitability of the data extraction form. Modifications to the data extraction form will be made as necessary and additional categories may be identified during data extraction. Any modifications made to the tool will be carefully noted and disclosed as part of our audit trail. In instances where essential data for extraction is not readily available within the published paper, authors of the respective publications will be contacted for clarification.

## Stage 5: Collating, summarizing and reporting the results

Scoping reviews, differing from systematic reviews, generally do not evaluate the methodological quality or bias risk of the studies included, nor do they perform data synthesis like meta-analyses. Instead, they offer a descriptive overview of the studies encompassed.^20^ Our approach involves both numerical and narrative summarization of various aspects of the included studies. These aspects include the year of publication, geographical location, clinical setting, participants, and study design. This analysis aims to map how silence is described in clinical interactions between patients and physicians.

Qualitative data will be analyzed using thematic analysis.^27^This will involve coding extracted data and grouping it iteratively to identify patterns of shared meaning in the data extracted. This will be complimented by drawing on our experiences as author group of family physicians (MK, CS, DB), palliative care doctors (SM, EmL) and medical educationalists (MB) working across different health contexts. We anticipate the need for regular team meetings to facilitate data charting and analysis.^24^

## Stage 6: Consultation

In line with Levac et al.’s recommendation, we will integrate consultation as a component of our planned scoping review.^24^ The outcomes of this review will be shared through presentations to palliative care physicians, ensuring a comprehensive exploration and understanding of silence’s role in clinical healthcare settings. This consultation phase will gather insights into our initial findings and their significance, explore potential applications and dissemination strategies, and identify areas requiring further research. Through this collaborative approach, we will facilitate discussions on implications and practical insights e.g. to improve communication skills training. Engaging in conversations with physicians will offer valuable insights into the practical implications of the research, enriching our skill set by incorporating diverse perspectives into the research process.

## Limitations

Whilst we will do our best to identify all relevant literature, it may be that our search strategy may miss some studies. Additionally, our search is restricted to English language studies across 5 databases. This protocol is restricted to physicians and physicians in training. Whilst understanding silence across health care professionals would be informative, the decision to restrict our search to physicians is based on a mix of pragmatic limitations and to focus our findings relative to our expertise in medical education. We anticipate study heterogeneity, which may limit the type of analysis possible.

## Discussion

This protocol presents the methodological framework and approach we will employ to identify and map the existing evidence regarding the experiences of physicians with silence in clinical settings. By identifying, analyzing, and synthesizing existing literature on this topic, the review will offer insights into the various roles and functions of silence in physician-patient interactions. The review will help recognize the diverse roles that silence plays in clinical consultations.

These roles encompass diverse aspects such as invitational silence, emotional acknowledgment, non-productive silence, and hostile silence, among others. Understanding these roles holds the potential to enhance the communication skills of healthcare professionals, enriching patient care experiences, improving their quality of life, and fostering a safe and comfortable environment within healthcare settings. Such insights can inform the development of tailored educational initiatives aimed at augmenting physicians’ proficiency in communication. Through dissemination via peer-reviewed presentations and publications, the findings of this scoping review will contribute to ongoing dialogues on optimizing doctor-patient communication and refining healthcare delivery practices.

### Authors’ contributions

Conceptualization: MK, SR, MB, CS, Methodology: all, Writing: MK and SR wrote the initial draft, this was reviewed and edited by all members of the team, who approved submission of the final protocol.

## Supporting Information

PRISMA-P

## Data Availability Statement

No datasets were generated or analyzed during the current study. All relevant data from this study will be made available upon study completion.

## Data Availability

This is a protocol for a literature review. All data reported in our literature review will already be in the public domain.

